# Adhering to dietary guidelines does not yield flavanol intake levels associated with beneficial cardiovascular effects

**DOI:** 10.64898/2026.02.24.26346949

**Authors:** Javier I. Ottaviani, John W. Erdman, Francene M. Steinberg, JoAnn E. Manson, Howard D. Sesso, Hagen Schroeter, Gunter G. C. Kuhnle

## Abstract

Outcomes from the COSMOS trial have reinforced the notion of flavanols as important plant-derived bioactives contributing to cardiovascular health. As discussions continue on whether specific dietary reference values for flavanols are warranted, it is possible that existing dietary guidelines emphasizing fruits and vegetables already yield sufficient flavanol intake levels. If this were the case, developing flavanol specific dietary reference values might be unnecessary. This study therefore aimed at assessing whether adherence to dietary recommendations for fruit and vegetable intake and overall diet quality achieves flavanol intake levels of 500 mg/day, the amount proven to mediate cardiovascular benefits in the COSMOS trial. Flavanol intake was objectively evaluated using two validated and complementary biomarkers, 5-(3□,4□-dihydroxyphenyl)-γ-valerolactone metabolites (gVLM_B_) and structurally related (–)-epicatechin metabolites (SREM_B_), in two geographically distinct studies: COSMOS (US; n=6,509) and EPIC-Norfolk (UK; n=24,154). The results showed that higher fruit and vegetable intakes and diet quality (assessed via the alternative healthy eating index–aHEI) were associated with increased flavanol intake in COSMOS. Nevertheless, fewer than 25% of participants meeting dietary guidelines achieved an estimated flavanol intake of ≥500 mg/day. Similar findings were observed in EPIC-Norfolk as well as through flavanol intake simulations considering fruits and vegetables commonly consumed in the US diet. In conclusion, adherence to existing dietary guidelines does not yield flavanol intake levels comparable to those shown to provide cardiovascular benefits in COSMOS. Thus, specific dietary reference values for flavanols may still be necessary if aiming to increase the intake of these dietary compounds.

**Graphical abstract:** 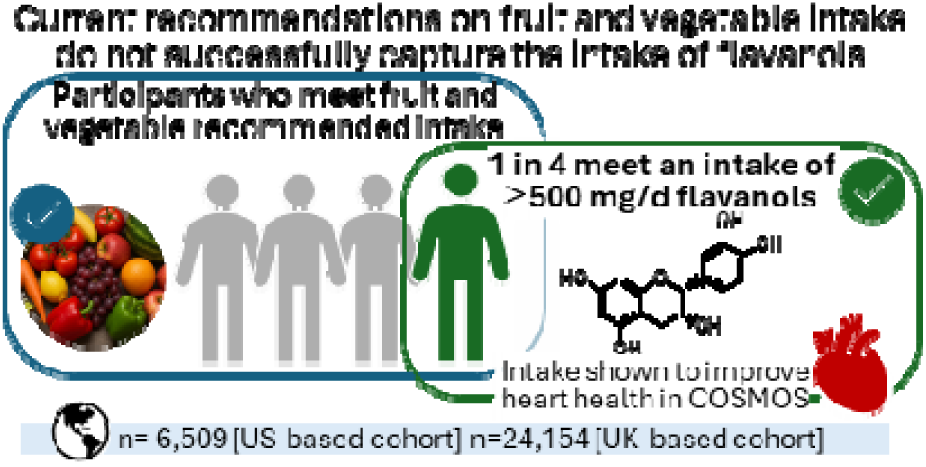

## Introduction

Flavanols are a distinct group of plant-derived polyphenolic bioactives (Ottaviani et al., 2018; Raman et al., 2019). Evidence from the COSMOS trial, the largest randomized controlled study on polyphenols to date, demonstrated that an intake of 500 mg/day of flavanols significantly reduced cardiovascular disease (CVD) mortality by 27% in intention-to-treat analyses, total cardiovascular events by 15% in per-protocol analyses and major cardiovascular events by 16% as a post-hoc endpoint analysed in intention-to-treat analyses in healthy older adults (Sesso et al., 2022). These findings have strengthened the view of flavanols as important dietary compounds for cardiovascular health, which have fuelled ongoing debates on whether specific dietary recommendations for flavanols and other bioactives should be established (Lupton et al., 2014; Erdman, 2023; Gaine et al., 2013; Yates et al., 2021). Recently, an expert scientific panel commissioned by the Academy of Nutrition and Dietetics in the United States proposed an intake of 400–600 mg/day for cardiometabolic health (Crowe-White et al., 2022). However, official dietary recommendations for flavanols have not yet been considered. Because fruits and vegetables are major sources of flavanols, and fruits and vegetables are emphasized in current dietary guidelines, it is plausible that adherence to current dietary guidelines could already deliver sufficient flavanol intake levels. Yet no data currently demonstrate whether following recommended intakes of fruits, vegetables, and overall healthy dietary patterns achieves flavanol levels associated with the cardiovascular benefits as shown in COSMOS trial. Addressing this point will provide valuable insights to determine the necessity of developing specific dietary recommendations for flavanols.

Flavanols are found in fruits like pome fruits, berries and stone fruit, vegetables like pinto, kidney and fava beans as well as other products like tea and cocoa-derived products. Previous studies have relied on self-reported dietary assessments to assess flavanol intake. However, these tools have shown significant limitations (Kuhnle, 2018; Ottaviani et al., 2024). In contrast, validated nutritional biomarkers, such as 5-(3□,4□-dihydroxyphenyl)-γ-valerolactone metabolites (gVLM_B_) and structurally related (–)-epicatechin metabolites (SREM_B_), offer more accurate estimates of flavanol intake (Ottaviani et al., 2019; Ottaviani et al., 2018). Furthermore, gVLM_B_ and SREM_B_ can assess the intake of those types of flavanols present in fruits and vegetables as well as those tested in the COSMOS trial (Figure 1). Comprehensive dietary and biomarker data are also available for two large studies, COSMOS (U.S.) and EPIC Norfolk (U.K.), making these cohorts ideally suited to examine the link between dietary recommendations and flavanol intake.

**Figure 1:**
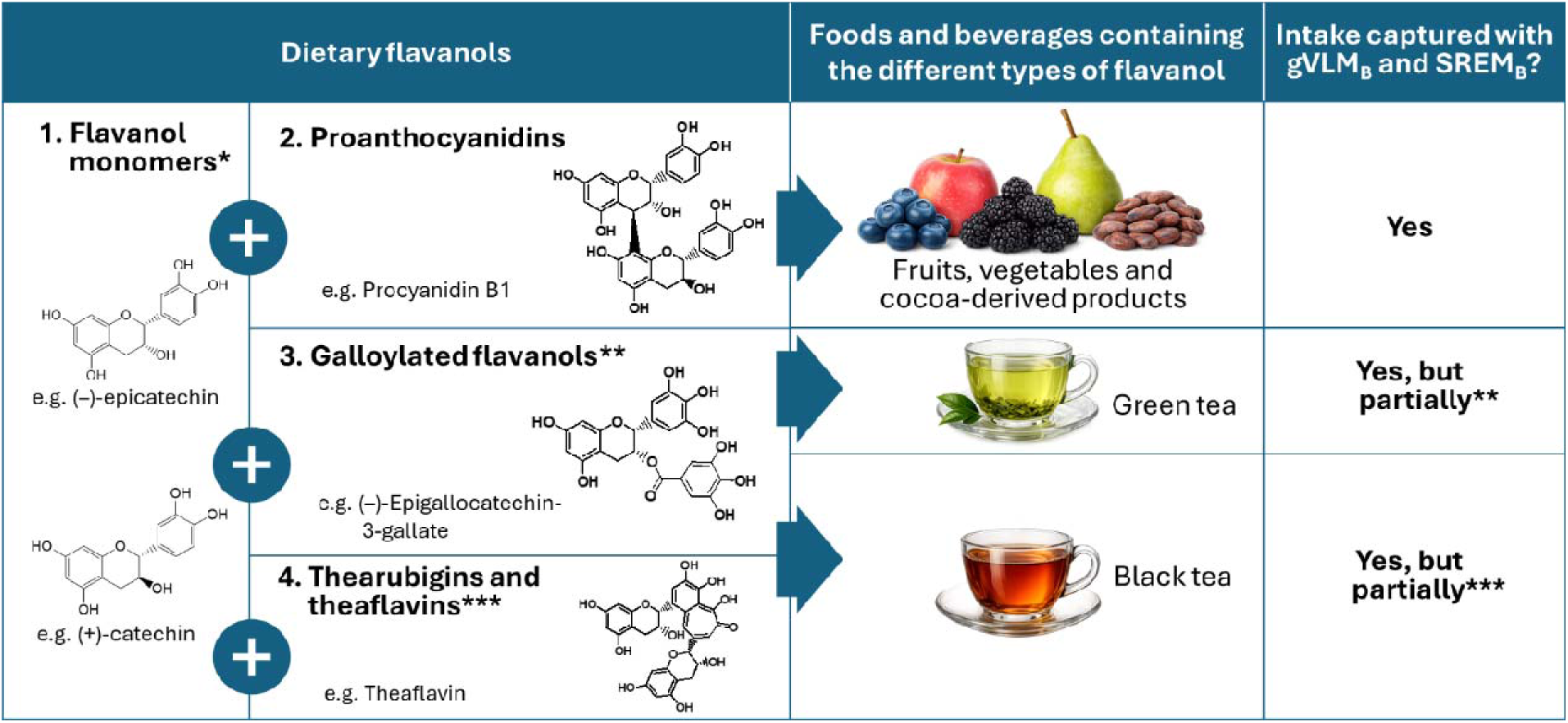
Main group of dietary flavanols present in different flavanol-containing food and beverages in the diet and contribution to flavanol biomarkers. *(–)-Epicatechin is the only flavanol monomer that contributes to SREM_B_ levels; ** Epicatechin-3-gallate and catechin-3-gallate are the galloylated flavanols that give rise to gVLM_B_ levels after intake; ***Thearubigins and theaflavins are exclusively present in black tea and the intake of these compounds does not give rise to gVLM_B_ and SREM_B_ levels. gVLM_B_: 5-(3□,4□-dihydroxyphenyl)-γ-valerolactone metabolites; SREM_B_: structurally related (–)-epicatechin metabolites

Here, we tested whether adhering to recommended levels of fruit and vegetable intake and an overall healthy dietary pattern results in flavanol intakes of at least 500 mg/d. This threshold reflects the intake shown to confer benefits in the COSMOS trial (Sesso et al., 2022) and also represents the average intake recommended by the guidelines commissioned by the Academy of Nutrition and Dietetics (Crowe-White et al., 2022). We used more recently collected data and assays in COSMOS to develop and assess this hypothesis and EPIC Norfolk to assess replication in an independent, population-based sample with differing dietary patterns and temporal context.

## Methodology

### Study design

#### COSMOS

The COSMOS trial (NCT02422745) is a randomized, double-blind, placebo-controlled, 2-by-2 factorial trial of a daily cocoa extract as a source of flavanols and a daily multivitamin for prevention of respective cardiovascular disease (CVD) and cancer among 21,442 US adults (12,666 women aged ≥65 y and 8776 men aged ≥60 y), free of major CVD and recently diagnosed cancer. COSMOS recruited participants from three main sources, including the Women’s Health Imitative (WHI), non-randomized recruitment respondents for the VITamin D and OmegA-3 TriaL (NCT01169259), and those identified through mass mailings and media efforts. Contact was made with nearly three million prospective participants. There were 21,442 participants ultimately enrolled and randomized into COSMOS. For the aspects relevant to this study, we conducted a post hoc analysis of data collected before randomization into COSMOS (i.e. before participants received any of the interventions). Fruit and vegetable intake as well as diet quality assessments were derived from a semi-quantitative food frequency questionnaire that participants completed during the run-in phase of the study (Feskanich et al., 1993; Rist et al., 2022). In addition, a subgroup of participants (n=6,509) provided spot urine samples also during the run-in phase of the study in which flavanol biomarkers were measured. Further details of the protocol and main findings of the study were previously published (Sesso et al., 2022). All participants provided written informed consent, and study approvals were obtained by the Institutional Review Board (IRB) at Mass General Brigham.

#### EPIC Norfolk

European Prospective Investigation into Cancer and Nutrition (EPIC)-Norfolk is an observational cohort study in Norfolk, UK. Between 1993 and 1997, 30,447 women and men aged between 40 and 79 years were recruited for the Norfolk cohort of the EPIC study, and 25,639 attended a health examination (Day et al., 1999). Diet was assessed by 7-day diary (7DD), whereby the first day of the diary was completed as a 24-hr recall (24HDR) with a trained interviewer and the remainder completed during subsequent days. Diary data were entered, checked, and calculated using the in-house dietary assessment software DINER (Data into Nutrients for Epidemiological Research) and DINERMO (Welch et al., 2001). Spot urine samples were collected during the health examination and stored at –20°C until analysis in which flavanol biomarkers were measured (Ottaviani et al., 2020). In addition, non-fasting blood samples were taken by venipuncture and stored in serum tubes in liquid nitrogen. Plasma vitamin C was measured using a fluorometric assay as described previously (Khaw et al., 2001). The study was approved by the Norwich Local Research Ethics Committee, all participants gave written, informed consent, and all methods were carried out in accordance with relevant guidelines and regulations.

#### Flavanol biomarkers

Flavanol biomarkers assessed in this study included the urinary levels of 5-(3□,4□-dihydroxyphenyl)-γ-valerolactone metabolites (gVLM_B_) and of the structurally related (–)-epicatechin metabolites (SREM_B_) (Ottaviani et al., 2019, 2018). While gVLM_B_ informs on the intake of flavanols in general, SREM is a specific biomarker of the intake of (–)-epicatechin, one of the main bioactive flavanol compounds and present within the 500 mg/d of flavanols tested in COSMOS. As such, gVLM_B_ and SREM_B_ provide information on the intake of the cocoa flavanols tested in COSMOS as well as the flavanols present in fruits and vegetables (Figure 1). While gVLM_B_ and SREM_B_ also provides information on the intake of flavanols from tea, one of the main dietary sources of flavanols, these biomarkers reflect the intake of those flavanols in tea that are also found in fruits and vegetables (Figure 1). Flavanols that are specific to tea, specially theaflavins and thearubigins derived from black tea processing, and absent in fruits and vegetables, do not influence gVLM_B_ and SREM_B_ levels (Ottaviani et al., 2019; Ottaviani et al., 2018).

SREM_B_ and gVLM_B_ have different systemic half-lives (Ottaviani et al., 2016), thus a combination of both biomarkers allows capturing different periods after flavanol intake. Adherence with flavanol intake consistent with the intake of 500 mg/d was estimated by using a combination of SREM_B_ and gVLM_B_ concentrations as described previously (Ottaviani et al., 2025). In brief, we calculated threshold values for the concentration of SREM_B_ and gVLM_B_ that could be expected after the intake of 500 mg of flavanols from a dose-escalation study that was part of the validation of SREM_B_ and gVLM_B_ as biomarkers (Ottaviani et al., 2019, 2018). In this manner, participants with SREM_B_ urinary concentration above 7.77 µM or a gVLM_B_ urinary concentration above 18.21 µM were considered to have an intake of flavanols of at least 500 mg/d.

gVLM_B_ corresponded to the sum of 5-(4□-hydroxyphenyl)-γ-valerolactone-3′-sulafte and 5-(4□-hydroxyphenyl)-γ-valerolactone-3′-glucuronide meanwhile SREM_B_ corresponded to the sum of (–)-epicatechin-3′-glucuronide, (–)-epicatechin-3′-sulafte and 3′-O-methyl-(–)-epicatechin-5-sulfate. These metabolites were quantified using validated LC-MS methods using authentic and isotopically labelled standards (Ottaviani et al., 2020). Unadjusted biomarker concentrations were used as urinary creatinine is associated with CVD risk (Ottaviani et al., 2023) and adjusting by specific gravity did not materially change biomarker levels.

### Diet quality assessment

#### Intake of fruit and vegetable

For COSMOS, we determined total fruit and vegetable intake (as servings per day) using data from a semi-quantitative food frequency questionnaire completed by participants during the run-in phase of the study prior to randomization (Feskanich et al., 1993; Rist et al., 2022). For EPIC Norfolk, we determined total fruit and vegetable intake (as g/d) from 7-day food diaries. In addition, vitamin C in plasma was assessed as a biomarker of fruit and vegetable intake (Bingham et al., 2008). Given the importance of tea as a dietary source of flavanols, tea intake was also assessed in both COSMOS and EPIC Norfolk, using the same corresponding dietary assessment methods for fruit and vegetable intake.

#### Diet quality

Diet quality was assessed with the alternative healthy eating index (aHEI). aHEI is an alternative index to the Healthy Eating Index that reflects earlier iterations of the Dietary Guidelines for Americans, including a modification to better predict impact on chronic diseases, including CVD (McCullough et al., 2002). In addition to total fruit and total vegetable intake, aHEI also included the assessment of nuts that could represent additional sources of flavanol intake (Bhagwat et al., 2015; Rothwell et al., 2013). aHEI was calculated for COSMOS but not for EPIC-Norfolk. Diet scores range from 0 to 87.5, in which 87.5 indicates perfect adherence.

### Simulation of flavanol intake from fruit and vegetable consumption

Flavanol intake from fruit and vegetable consumption was simulated using food composition data from Phenol-Explorer (PhenolExplorer) and 2017 NHANES data on fruit and vegetable consumption patterns. We evaluated varying numbers of daily portions (using sizes based on 21 CFR § 101.12) and different food selection strategies: (1) random (unweighted), (2) weighted by US consumption frequency (NHANES 2017), and (3) weighted by flavanol content. For each combination of portion number, selection strategy, and sampling scheme (with or without replacement), we conducted 10,000 Monte Carlo simulations. At each iteration, flavanol content of a selected food item was drawn from a uniform distribution defined by the reported minimum and maximum concentrations in Phenol-Explorer, based on the sum of individual flavanols that give rise to flavanol biomarkers, including (–)-epicatechin, (+)-catechin, epicatechin 3-O-gallate, catechin 3-O-gallate, and procyanidins from dimers up to decamers (Figure 1). The simulations used Full data, and code can be found here: https://gitlab.act.reading.ac.uk/xb901875/fruit_vegetable_flavanols

### Statistical analysis

All analyses were conducted with R version 4.3 in RStudio 2024.09.0. Tables were created using the table one package (Yoshida and Bohn, 2015), regression analyses using the rms package (Harrell Jr, 2017) and graphics using ggplot2 (Wickham, 2009). Dietary data were log2 transformed before analysis. lrm was used to conduct logistic regressions with “meeting a biomarker-estimated flavanol intake of at least 500 mg/d” as dependent variable. Models were adjusted by age and sex, using restricted cubic splines for all continuous variables. Fruit and vegetable intakes were divided into quartiles using the cut function. For the aHEI score, ties prevented even distribution into quartiles. We added minimal random jitter (±0.0001) to break ties, repeating this process 1000 times and averaging results across iterations to avoid dependence on any single arbitrary tie-breaking assignment. Missing values were excluded from the analysis (White and Royston, 2009).

## Results

### Study population

Biomarker data were available for n=6,509 COSMOS participants and n=24,154 EPIC Norfolk participants (Table 1). In general, participants in COSMOS were older, had a slightly higher proportion of males, and a higher BMI compared to participants in EPIC Norfolk; EPIC Norfolk was less ethnically diverse. As intake of food items, including fruits and vegetables, was reported in different units in COSMOS (i.e. servings/day) and EPIC Norfolk (i.e. g/day), values were not directly compared between cohorts except for tea, which was consumed at much higher levels in EPIC Norfolk (773 ± 530 g/d) than COSMOS (134 ± 261 g/d). Concentrations of gVLM_B_, the more general biomarker of flavanol intake, were not different between studies (Table 1, and Suppl. Fig 1). However, SREM_B_ levels, the specific biomarker of (–)-epicatechin intake, were higher in participants in EPIC Norfolk compared to those in COSMOS (Table 1, and Suppl. Fig 1). Data from both biomarkers were combined to estimate the proportion of participants meeting an intake of at least 500 mg/d of flavanols (Ottaviani et al., 2025). The percentage of the population that met the biomarker-estimated flavanol intake of at least 500 mg/d was 19.2% and 17.9 % in COSMOS and EPIC, respectively (Table 1). In both studies, men were more likely to meet a flavanol intake of at least 500 mg/d (OR 1.36 (95% CI 1.29; 1.45); adjusted by age, BMI and recruitment cohort). In contrast, older participants of COSMOS were more likely to meet a flavanol intake of at least 500 mg/d (OR 1.21 (1.06; 1.39); 65 vs 75 years; adjusted by sex and BMI), while this was reversed in EPIC Norfolk (OR 0.91 (0.83; 0.99) 65 vs 75 years; adjusted by sex and BMI). Likewise, normal-weight participants of COSMOS were more likely to meet a flavanol intake of at least 500 mg/d (OR 0.92 (0.85; 0.98)) compared with obese participants, while the opposite was the case in EPIC (OR 1.12 (1.07; 1.17); Supplemental Table 1).

**Table 1:** Characteristics of study population in COSMOS (US) and EPIC Norfolk (UK). Data are expressed as mean ± SD, unless otherwise stated.

|  | COSMOS | EPIC Norfolk | p<sup>1</sup> |
| --- | --- | --- | --- |
| Participants (n) | 6,509 | 24,154 |  |
| Age (years) | 71.0 ± 6.3 | 58.6 ± 9.3 | <0.001 |
| Male (n) | 3,178 (48.8%) | 10,879 (45.0%) | <0.001 |
| BMI (kg/m<sup>2</sup>) | 27.6 ± 5.3 | 26.3 ± 3.9 | <0.001 |
| Fruit & vegetable intake (COSMOS, servings/day; EPIC Norfolk, g/day)<sup>2</sup> | 5.71 ± 4.13 | 574 ± 341 | n.d. |
| Fruit intake (COSMOS, servings/day; EPIC Norfolk, g/day)<sup>2</sup> | 2.27 ± 1.96 | 161 ± 133 | n.d. |
| Vegetable intake (COSMOS, servings/day; EPIC Norfolk, g/day)<sup>2</sup> | 3.45 ± 2.82 | 121 ± 74 | n.d. |
| Tea intake (g/day)<sup>2</sup> | 134 ± 261 | 773 ± 530 | <0.001 |
| Plasma vitamin C (μmol/L) | n.d.<sup>3</sup> | 53.5 ± 20.4 | n.d. |
| aHEI Score | 42.56 ± 10.82 | n.d.<sup>3</sup> | n.d. |
| gVLM<sub>B</sub> (μmol)<sup>2</sup> | 3.30 [0.78, 11.00] | 3.24 [0.83, 10.47] | 0.126 |
| SREM<sub>B</sub> (μmol)<sup>2</sup> | 0.47 [0.10, 1.88] | 0.87 [0.23, 2.38] | <0.001 |
| Participants meeting a biomarker-estimated flavanol intake of at least 500 mg/d (n) | 1248 (19.2%) | 4317 (17.9%) | 0.016 |

<sup>1</sup> Test for difference is between the two cohorts determined by t-test, except for gVLM<sub>B</sub> and SREM<sub>B</sub> that were compared using Fischer's test. <sup>2</sup> Data are expressed as median and IQR. <sup>3</sup> Not determined. gVLM<sub>B</sub>: 5-(3,4-dihydroxyphenyl)- γ-valerolactone metabolites; SREM<sub>B</sub>: structurally related (–)-epicatechin metabolites

### Association between diet quality and biomarker-estimated flavanol intake in COSMOS

In order to investigate whether participants that adhere to current dietary guidelines have a higher flavanol intake, we divided participants into quartiles based on fruit and vegetable intake and diet quality assessed as aHEI score (Table 2). Participants in the top quartile of fruit and vegetable intake met and often surpassed the 5 servings/day recommendation by the Dietary Guidelines for Americans (U.S. Department of Agriculture and U. S. Department of Health and Human Services, 2020). Similarly, participants in the top quartile of aHEI showed scores between 58% to 93% of maximal score, suggesting that participants in the highest quartile of aHEI presented high adherence to a healthy dietary pattern. However, despite meeting these recommendations and following a healthy dietary pattern, only 21% of participants consumed at least 500 mg/d of flavanols (Table 2). These figures were not substantially higher than the 19.2% of participants with a flavanol intake of at least 500 mg/d determined for the entire COSMOS cohort (Table 1). As tea can be an important source of flavanols (Vogiatzoglou et al., 2015), we also included tea in our analyses. COSMOS participants reported relatively low tea consumption, with over one-third reporting no intake. There was no significant difference in the proportion of participants meeting the 500 mg/day flavanol threshold between high and low tea consumers (Table 2).

**Table 2:** Number of COSMOS^2^ participants meeting a biomarker-estimated flavanol intake of at least 500 mg/d across different quartiles of fruit and vegetable intake, tea intake and diet quality. Data are shown as mean ± SD.

| **Dietary assessment** | **Quartile 1** | **Quartile 2** | **Quartile 3** | **Quartile 4** | **p<sup>1</sup>** |
| --- | --- | --- | --- | --- | --- |
| **Fruit & vegetable intake (serving/d)** | **0.1 – 3.4** | **3.5 – 5** | **5 – 7.1** | **7.1 – 121<sup>5</sup>** |  |
| COSMOS participants (n) | 1549 | 1547 | 1548 | 1548 |  |
| Age (years) | 69.8 ± 5.9 | 70.5 ± 5.9 | 71.6 ± 6.4 | 72.2 ± 6.7 | <0.001 |
| Male (n) | 831 (54%) | 770 (50%) | 704 (46%) | 632 (41%) | <0.001 |
| Participants meeting a biomarker-estimated flavanol intake of at least 500 mg/d (n) | 265 (17%) | 298 (19%) | 297 (19%) | 322 (21%) | 0.075 |
| **Fruit intake (serving/day)** | **0 – 1.1** | **1.1 – 1.9** | **1.9 – 2.9** | **2.9 – 36.1<sup>5</sup>** |  |
| COSMOS participants (n) | 1554 | 1553 | 1539 | 1548 |  |
| Age (years) | 69.7 ± 5.7 | 70.7 ± 6.1 | 71.4 ± 6.2 | 72.32 ± 6.9 | <0.001 |
| Male (n) | 818 (53%) | 740 (48%) | 707 (46%) | 674 (44%) | <0.001 |
| Participants meeting a biomarker-estimated flavanol intake of at least 500 mg/d (n) | 267 (17%) | 301 (19%) | 308 (20%) | 306 (20%) | 0.167 |
| **Vegetable intake (serving/day)** | **0 – 1.9** | **1.9 – 2.9** | **2.9 – 4.3** | **4.3 – 85.1<sup>5</sup>** |  |
| COSMOS participants (n) | 1553 | 1555 | 1550 | 1552 |  |
| Age (yrs) | 70.2 ± 6.2 | 70.4 ± 6.0 | 71.6 ± 6.4 | 71.9 ± 6.4 | <0.001 |
| Male (n) | 845 (54%) | 800 (51%) | 684 (44%) | 621 (40%) | <0.001 |
| Participants meeting a biomarker-estimated flavanol intake of at least 500 mg/d (n) | 287 (19%) | 300 (19%) | 297 (19%) | 305 (20%) | 0.868 |
| **Tea intake (servings/day)<sup>3</sup>** | **0 – 0.1 (bottom half)** |  | **0.1 – 14 (top half)** |  |  |
| COSMOS participants (n) | 3433 |  | 2732 |  |  |
| Age (yrs) | 70.8 ± 6.4 | | 71.3 ± 6.1 | | 0.001 |
| Male (n) | 1872 (55%) |  | 1054 (39%) |  | <0.001 |
| Participants meeting a biomarker-estimated flavanol intake of at least 500 mg/d (n) | 651 (20%) |  | 530 (20%) |  | 0.689 |
| **aHEI<sup>4</sup> score** | **12.5 – 34.5** | **34.5 – 42.5** | **42.5 – 50.5** | **50.5 – 81.5** |  |
| COSMOS participants (n) | 1481 | 1480 | 1480 | 1480 |  |
| Age (years) | 70.2 ± 6.1 | 71.0 ± 6.4 | 71.2 ± 6.3 | 71.7 ± 6.4 | <0.001 |
| Male (n) | 793 (54%) | 756 (51%) | 756 (47%) | 696 (38%) | <0.001 |
| Participants meeting a biomarker-estimated flavanol intake of at least 500 mg/d (n) | 240 (16%) | 281 (19%) | 281 (19%) | 283 (22%) | <0.001 |

<sup>1</sup> Test for difference is between quartiles was determined by ANOVA except for aHEI that was determined using a bootstrapping approach with meta-regression of results. Differences were determined using meta-regression analyses. <sup>2</sup>COocoa Supplement and Multivitamin Outcomes Study. <sup>3</sup>Almost 50% of COSMOS participants did not consume any tea and a division into quartiles was therefore not possible and data presented correspond to top and bottom half of the population. <sup>4</sup>aHEI: Alternative Healthy eating Index. <sup>5</sup>Fewer than 1% of participants reported unrealistically high fruit and vegetable intake (99<sup>th</sup> percentile: 8, 11 and 18 servings/per day of fruits, vegetables and fruit and vegetables.

We initially compared unadjusted quartiles of fruit, vegetable, fruit and vegetable intake, and aHEI score in relation to meeting the flavanol intake threshold of 500 mg/day. To investigate whether these association held after accounting for potential confounders, we calculated odds ratios (OR) adjusted for age, sex, and BMI. Participants in the highest aHEI quartile had 26% higher odds of meeting the 500 mg/day threshold compared to those in the lowest quartile (Table 3). For all other markers of healthy diet, no significant OR differences were observed (Table 3). Additional analyses showed that fruit intake, aHEI and tea intake were weakly but significantly associated with SREM_B_ and gVLM_B_ concentrations (Supplemental Table 2).

**Table 3:** Association between food intake and diet quality score and meeting a biomarker-estimated flavanol intake of at least 500 mg/d in COSMOS. Odds ratio (OR) comparing top versus bottom quartiles of diet quality score among COSMOS participants. Data was log2 transformed and regression analysis was adjusted for age, sex, and BMI.

| **<u>Dietary assessment</u>** | **OR (95% CI)** | **p** |
| --- | --- | --- |
| Fruit & vegetable intake | 1.10 (1.01; 1.19) | 0.06 |
| Vegetable* intake | 1.04 (0.95; 1.14) | 0.44 |
| Fruit* intake | 1.05 (0.96; 1.16) | 0.22 |
| Tea intake | 0.99 (0.93; 1.06) | 0.30 |
| aHEI score | 1.26 (1.14; 1.39) | < 0.001 |

\*Model additionally adjusted for fruit and vegetable intake respectively. P-value for Wald test

### Association between diet quality and biomarker-estimated flavanol intake in EPIC-Norfolk

In order to investigate whether the findings obtained in COSMOS participants (Table 2 and 3) are also observed in a different cohort and of a different geographical region, the association between fruit and vegetable intake and odds to meet a flavanol intake of at least 500 mg/d was assessed in participants of EPIC Norfolk (Table 4). Participants in the top quartile of fruit and vegetable intake indeed showed intakes meeting or surpassing the amounts recommended by the 5-a-day recommendation (Public Health England, 2016; U.S. Department of Agriculture and U. S. Department of Health and Human Services, 2020)UK Eatwell Guide, Public Health England). Instead of using aHEI, which was not available, we used plasma vitamin C as surrogate marker of a healthy diet as described previously (Ottaviani, 2020). Only 16% of participants in the top quartile presented a flavanol intake of at least 500 mg/d (Table 4). Similar to COSMOS, adhering to a higher fruit and vegetable intake did not result in higher odds to meet a flavanol intake of at least 500 mg/d (Table 4). Similar findings were observed when comparing number of participants meeting a flavanol intake of at least 500 mg/d between top and bottom quartiles of fruit intake and vegetable intake (Table 4). Differences in the number of participants meeting a flavanol intake of at least 500 mg/d were more pronounced between top and bottom quartiles of vitamin C levels (Table 4). Consistently, there was a modest inverse relationship between fruit and vegetable intake and the odds of meeting a flavanol intake of at least 500 mg/d in EPIC Norfolk participants (Table 5). Similar findings were observed when assessing fruit intake, vegetable intake, and plasma vitamin C (Table 5). These results show that those not following dietary recommendations generally were more likely to have a flavanol intake of 500 mg/d as confirmed by the adjusted ORs (Table 5). In contrast, those with high tea intake were more likely to attain 500 mg/d of flavanol intake, albeit even in the highest quartile of tea intake, only 19% of participant achieved this intake (Table 5).

**Table 4:** Number of EPIC Norfolk^2^ participants meeting a biomarker-estimated flavanol intake of at least 500 mg/d across different quartiles of fruit and vegetable intake, tea intake and vitamin C concentration. Data are shown as mean ± SD.

| **<u>Dietary assessment</u>** | **Quartile 1** | **Quartile 2** | **Quartile 3** | **Quartile 4** | **p<sup>1</sup>** |
| --- | --- | --- | --- | --- | --- |
| **Fruit & vegetable intake (g/d)** | 0 - 336.2 | 336.2 - 520.8 | 520.9 - 747.3 | 747.3 - 4762.8 |  |
| EPIC-Norfolk participants (n) | 6010 | 6009 | 6009 | 6010 |  |
| Age (years) | 57.84 (9.54) | 58.62 (9.35) | 58.99 (9.23) | 58.81 (8.86) | <0.001 |
| Male (n) | 3180 (52.9) | 2786 (46.4) | 2520 (41.9) | 2331 (38.8) | <0.001 |
| Participants meeting a biomarker-estimated flavanol intake of at least 500 mg/d (n) | 1196 (19.9) | 1104 (18.4) | 1029 (17.1) | 964 (16.0) | <0.001 |
| **Fruit intake (g/day)** | 0 - 63.4 | 63.4 - 136.6 | 136.6 - 226.6 | 226.6 - 2100 |  |
| EPIC-Norfolk participants (n) | 6010 | 6009 | 6009 | 6010 |  |
| Age (years) | 57.62 (9.41) | 58.54 (9.36) | 59.17 (9.18) | 58.93 (8.99) | <0.001 |
| Male (n) | 3358 (55.9) | 2688 (44.7) | 2500 (41.6) | 2271 (37.8) | <0.001 |
| Participants meeting a biomarker-estimated flavanol intake of at least 500 mg/d (n) | 1195 (19.9) | 1086 (18.1) | 1032 (17.2) | 980 (16.3) | <0.001 |
| **Vegetable intake (g/day)** | 0 - 72.2 | 72.2 - 109.7 | 109.7 - 157 | 157 - 1180.8 |  |
| EPIC-Norfolk participants (n) | 6010 | 6009 | 6009 | 6010 |  |
| Age (years) | 58.11 (9.67) | 58.88 (9.24) | 58.64 (9.19) | 58.62 (8.91) | <0.001 |
| Male (n) | 2847 (47.4) | 2712 (45.1) | 2619 (43.6) | 2639 (43.9) | <0.001 |
| Participants meeting a biomarker-estimated flavanol intake of at least 500 mg/d (n) | 1165 (19.4) | 1084 (18.0) | 1053 (17.5) | 991 (16.5) | <0.001 |
| **Tea intake (g/d)** | 0 – 417 | 418 – 731 | 731 – 1069 | 1069 – 6975 |  |
| EPIC-Norfolk participants (n) | 6016 | 6007 | 6006 | 6009 |  |
| Age (years) | 56.74 (8.99) | 59.61 (9.32) | 59.84 (9.29) | 58.08 (9.09) | <0.001 |
| Male (n) | 2694 (44.8) | 2552 (42.5) | 2599 (43.3) | 2972 (49.5) | <0.001 |
| Participants meeting a biomarker-estimated flavanol intake of at least 500 mg/d (n) | 899 (14.9) | 1084 (18.0) | 1161 (19.3) | 1149 (19.1) | <0.001 |
| **Vitamin C (μmol/L)** | 3.0-41 | 41.9 - 54 | 54.1 - 66 | 66.5 - 242 |  |
| EPIC-Norfolk participants (n) | 5474 | 5294 | 5324 | 5085 |  |
| Age (years) | 59.55 (9.36) | 58.43 (9.34) | 57.88 (9.03) | 58.02 (9.08) | <0.001 |
| Male (n) | 3476 (63.5) | 2799 (52.9) | 2048 (38.5) | 1295 (25.5) | <0.001 |
| Participants meeting a biomarker-estimated flavanol intake of at least 500 mg/d (n) | 1105 (20.2) | 969 (18.3) | 934 (17.5) | 807 (15.9) | <0.001 |

<sup>1</sup> Test for difference is between quartiles was determined by ANOVA.

**Table 5:** Association between food intake and diet quality score and meeting a biomarker-estimated flavanol intake of at least 500 mg/d in EPIC Norfolk. Odds ratio (OR) comparing top versus bottom quartiles of diet quality score among EPIC Norfolk participants. Data was log2 transformed and regression analysis was adjusted for age, sex, and BMI.

| **Dietary assessment** | **OR (95% CI)** | **p** |
| --- | --- | --- |
| Fruit & vegetable intake | 0.99 (0.98; 1.00) | <0.001 |
| Vegetable* intake | 0.99 (0.98; 1.00) | 0.006 |
| Fruit* intake | 1.00 (0.99; 1.01) | 0.005 |
| Tea intake | 1.14 (1.09; 1.20) | <0.001 |
| Vitamin C | 0.92 (0.88; 0.97) | 0.003 |

\*Model additionally adjusted for fruit and vegetable intake respectively. P-value for Wald test

Further analyses investigating the association between diet quality flavanol biomarkers were conducted (Supplemental Table 3). Intake of tea also showed slightly stronger (0.1<β <0.3) associations with both flavanol biomarkers, SREM_B_ and gVLM_B_ (Supplemental Table 3).

### Fruit and vegetable intake and flavanol intake simulation

The majority of study participants, even those with high fruit and vegetable intake, did not achieve a flavanol intake of at least 500 mg/d. To further evaluate these results but using a non-biomarker approach, we simulated the likelihood of ingesting 500 mg/d flavanol when consuming different numbers of portions of fruit and vegetable following different selection strategies (Figure 2). Consistent with our biomarker-based analysis, it is unlikely to reach 500 mg/d of flavanols when prioritising the intake of five portions of fruits and vegetables commonly consumed in the US (including bananas, apples, tomatoes, grapes, oranges, carrots, etc.; see Supplemental Table 4 for further details) or selected at random. When prioritising the consumption of fruits and vegetables with higher flavanol content, the probability of consuming 500 mg/d of flavanols increased but remained less than 50%.

**Figure 2:**
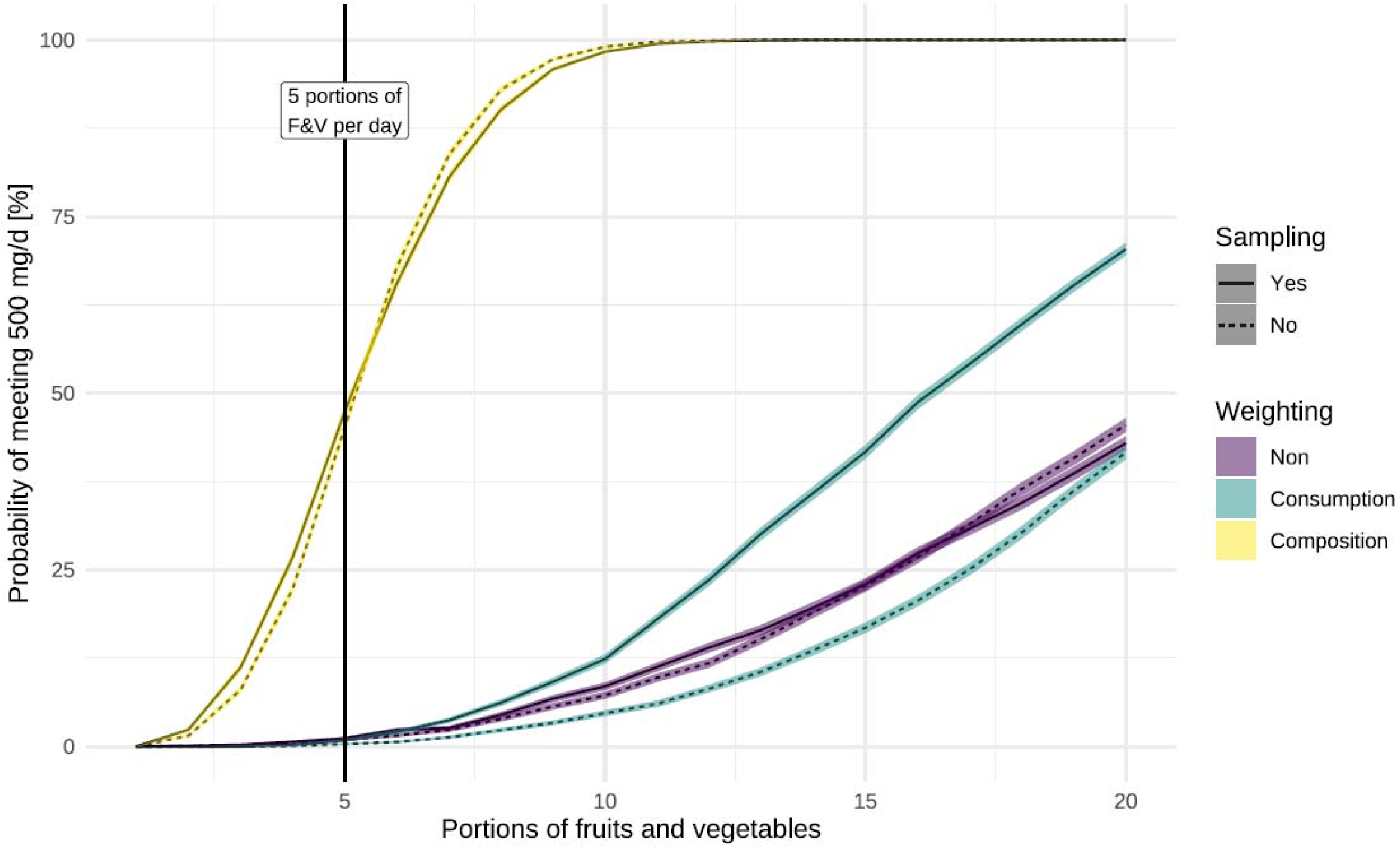
Probability of consuming at least 500 mg/d of flavanols with different numbers of servings of fruits and vegetables (F&V) using different selection methods, including weighing and sampling. F&V selection was either completely at random, weighted by typical consumption of F&V in the US or weighted by flavanol content in F&V. Fruits and vegetables sampling was done either with replacement (each F&V item can be selected more than once) or without replacement (reflecting the general recommendation of consuming different types of fruits and vegetables). Values shown are mean and 95% confidence interval of 10,000 simulations. Portion sizes for each F&V were based on 21 CFR § 101.12. At each iteration, flavanol content of a selected food item was drawn from a uniform distribution defined by the reported minimum and maximum concentrations in Phenol-Explorer.

## Discussion

In this study, we investigated whether following current dietary guidelines as measured by fruit and vegetable intake and aHEI sufficiently reflects flavanol intake of at least 500 mg/day, the amount found in COSMOS to significantly reduce cardiovascular mortality by 27% (Sesso et al., 2022). To overcome inherent limitations of self-reported dietary assessments to estimate flavanol intake (Ottaviani et al., 2024), we incorporated biomarker data to establish whether study participants consumed at least 500 mg/d of flavanols (Ottaviani et al., 2025). Our results show that even among participants who adhere to current recommendations for fruit and vegetable intake and healthy dietary patterns, less than 25% of participants achieved a biomarker-estimated flavanol intake of at least 500 mg/d (Table 2 and 4). Further, we found very modest differences in flavanol intake between participants with high versus low fruit and vegetable intake (Table 3 and 5). Thus, these results show that an adherence to current dietary guidelines does not ensure a level of flavanol consumption sufficient to expect beneficial cardiovascular effects as shown in the COSMOS trial (Sesso et al., 2022) and to meet the recommendations for flavanol intake commissioned by the Academy of Nutrition and Dietetics (Crowe-White et al., 2022).

The different iterations of the Dietary Guidelines for Americans were designed to meet dietary reference intake (DRI) for nutrients (U.S. Department of Agriculture and U. S. Department of Health and Human Services, 2020), not flavanols. Nevertheless, fruits and vegetables are among the main dietary sources of flavanols in the diet (Bhagwat et al., 2004, 2004; Rothwell et al., 2013) and are the focus of dietary guidelines. However, our data counter the assumption that meeting recommendations for fruit and vegetable intake will translate to an intake of flavanols of at least 500 mg/d based simulations considering the range of flavanol levels present in fruit and vegetables commonly consumed in the US (Figure 2). When prioritizing fruits and vegetables with high flavanol content, the probability to achieve such an intake moderately increased. However, increasing flavanol intake should not be achieved at the expense of decreasing the diversity of fruits and vegetables consumed. In this context, it is worth considering that flavanol content varies significantly within individual fruits and vegetables based on plant cultivars and breeds, climate, growing and harvest conditions (Wilkinson and Perring, 1961). The content of (–)-epicatechin even within the same apple variety can fluctuate more than 10-fold (Bhagwat et al., 2004, 2004; Rothwell et al., 2013), suggesting that the number of apples to meet an intake of 80 mg of (–)-epicatechin (which is the amount provided in the flavanol intervention tested in COSMOS) could vary from 2 to 29. Furthermore, procyanidins are flavanols that contribute to the astringency in fruits, that is usually identified as an undesired oral sensory quality that breeders may try to remove (Wu et al., 2022). Precisely, the development of specific dietary recommendations for flavanols may positively influence current agricultural practices and food selection methods to increase and optimize the content of flavanols in different plant foods and create an opportunity for food producers to develop varieties with higher flavanol content.

Flavanol intake in the two studies analyzed, COSMOS in the US and EPIC-Norfolk in the UK, showed that the odds of meeting an intake of 500 mg/d was similar despite differences in the dietary composition between these two countries and the time these studies were conducted. The levels of the more general biomarker of flavanols intake, gVLM_B_, was similar in both cohorts, while SREM_B_ showed higher levels in EPIC-Norfolk (Table 1 and Suppl. Figure 1). These results are probably due to the higher intake of tea in the UK (Table 1), which represents one of the main sources of (–)-epicatechin in the UK diet (Vogiatzoglou et al., 2014). Nevertheless, the proportion of participants meeting an intake of 500 mg/d of flavanols remained relatively low even within the top tea consumers in EPIC-Norfolk (Table 4 and 5). Given the relatively low intake of tea in COSMOS compared to that in EPIC, tea represents a valuable means to increase flavanol intake in the US. However, this should be done with certain considerations. For instance, Dietary Guidelines for Americans rather implicate tea as a potential means of incorporating added sugars, sweeteners, cream and caffeine (U.S. Department of Agriculture and U. S. Department of Health and Human Services, 2020). In addition, while black tea can contribute to the intake of flavanols such as theaflavins and thearubigins (Bhagwat et al., 2004, 2004; Rothwell et al., 2013), the contribution of these black tea-specific flavanols to the overall health benefits associated with the intake of the flavanols and procyanidins tested in COSMOS remains to be elucidated.

The key strength of this study is the use of validated biomarkers to objectively assess the intake of flavanols. Unlike self-reported dietary assessment methods, which are susceptible to recall bias and other inaccuracies (Kuhnle, 2018), biomarkers provide a more reliable measure. However, interindividual differences in the bioavailability of flavanols as well as sampling can influence the levels of these biomarkers (Ottaviani et al., 2025). These limitations have been discussed previously (Ottaviani et al., 2019, 2018), including those related to the combination of gVLM_B_ and SREM_B_ to assess the intake of at least 500 mg/d flavanols (Ottaviani et al., 2025). For our analyses, aHEI was used to assess diet quality as this represents previous iterations of the Dietary Guidelines for Americans. While other indexes could have been included in the analysis, simulations of flavanol intake showed that even the selection of five portions of fruits and vegetables high in flavanol did not result in flavanol intake of 500 mg/day or higher (Figure 2). Thus, it is expected that other dietary patterns recommended by the Dietary Guidelines for Americans such as the DASH and Mediterranean diet would not yield flavanol intakes of at least 500 mg/day. Another notable strength is the inclusion of two large studies of geographically different origin. It should be noted that COSMOS participants, who enrolled in a long-term randomized clinical trial, tended to have a considerably healthier dietary pattern compared to the general US public (Brickman et al., 2023), potentially leading to an overestimation of the amount of fruit and vegetable – and flavanol – consumption. Therefore, an even smaller proportion of older US adults likely meets the 500 mg/d threshold. Future research is therefore needed to evaluate flavanol intake within representative US and UK populations, to better understand the potential public health impact of increased flavanol consumption.

The results from this study also contribute to the general discussion of whether or not developing dietary reference values for bioactives is warranted (Lupton et al., 2014;Erdman, 2023; Gaine et al., 2013; Yates et al., 2021). As fruits and vegetables represent a main source of dietary bioactives like flavanols, it is plausible to expect that generic advice to increase fruit and vegetable consumption may not ensure optimal intake of specific bioactives. Such will be the case for other polyphenolic bioactives like anthocyanidins and flavanones as well as other bioactives such as carotenoids, which similar to flavanols, have a distribution in the diet that can vary across specific fruits and vegetables for its content. Considering the importance of diet in disease prevention and healthy aging, further debates and conversations on the development of DRIs or DRI-like values for bioactives offers stimulating opportunities for nutrition research. Perhaps rather than a direct pathway for inclusion of a quantitative DRI target for bioactives, a nuanced and qualitative approach to emphasizing rich food sources, or the development of foods with higher levels of specific bioactives, will be an indirect pathway to address bioactive intake.

## Conclusion

These results show that adherence to current dietary guidelines is not sufficient to address flavanol intake in amounts shown to significantly reduce the risk of cardiovascular death in the COSMOS trial. Hence, the development of specific dietary reference values for flavanols may still be necessary if aiming to increase the intake of these compounds from the diet and translate the health benefits related to the intake of these compounds to the general public. While the development of DRI□like values for flavanols and other bioactive compounds is not presently prioritized in the United States, other authoritative bodies may be able to advance such efforts in a shorter timeline. In this context, recent efforts to develop a framework for the development of recommended intakes for bioactives (Yates et al., 2021) as well as the guidelines for flavanol intake commissioned by the Academy of Nutrition and Dietetics (Crowe-White et al., 2022) represent valuable steps towards this goal.

## Author contributions

Designed research: JIO, GGCK, HS; performed research: JIO, GGCK; analyzed data: JIO, GGCK; wrote the paper: JIO, GGCK, JWE, FS, HS, HDS, JEM

## Data availability

Code is available from: https://gitlab.act.reading.ac.uk/xb901875/flavanol-dq

Data from COSMOS trial and associated documentation will be available to users only under a data-sharing agreement. Details on the availability of the study data to other investigators will be on our study website at: https://cosmostrial.org/.

EPIC Norfolk aims to make data and samples as widely available as possible whilst safeguarding the privacy of our participants, protecting confidential data and maintaining the reputations of our studies and participants aims. Information on how to request data from EPIC Norfolk can be found here: https://www.epic-norfolk.org.uk/for-researchers/data-sharing/data-requests/.

## Acknowledgements

We are deeply indebted to the 21L442 COSMOS participants for their steadfast and conscientious collaboration. We are grateful to all EPIC Norfolk participants who have been part of the project and the many members of the study teams at the University of Cambridge who have enabled this research.

## Supplementary information

**Supplemental Table 1:**
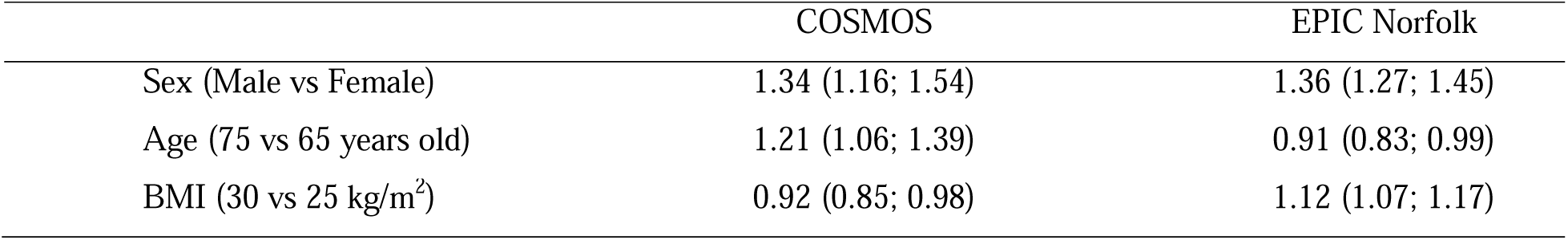
Odd ratio (OR) for meeting a biomarker-estimated flavanol intake of at least 500 mg/d in COSMOS and EPIC Norfolk, comparing sex, age and BMI using logistic regression analysis.

**Supplemental Table 2:**
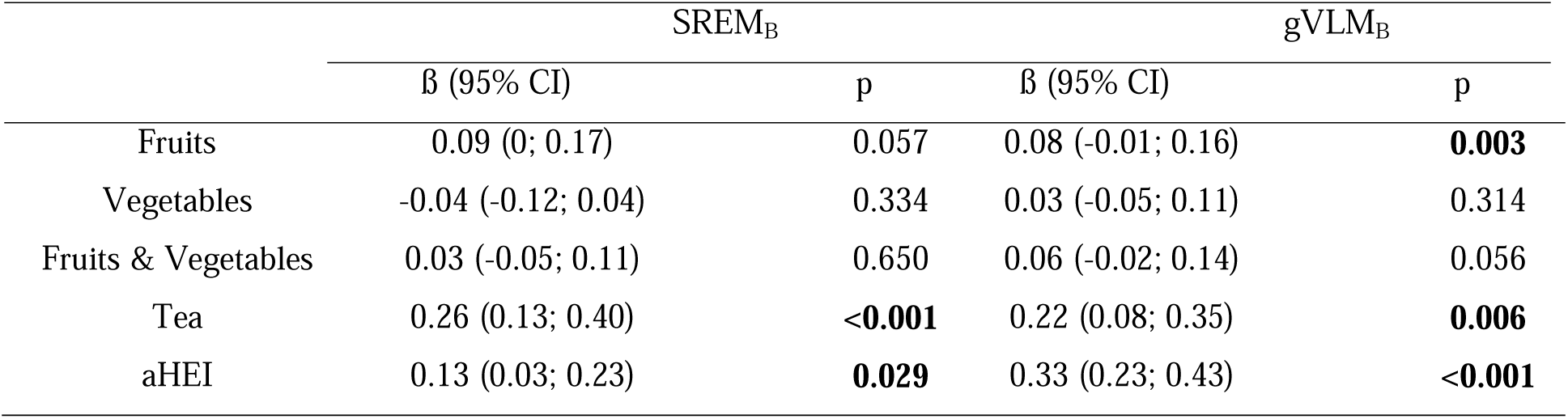
Association between markers of diet quality and urinary flavanol in COSMOS (difference in log2 biomarker between bottom and top quartile of respective marker, adjusted by age, sex and BMI; 95% confidence interval and Wald-test).

**Supplemental Table 3:** Association between markers of diet quality and urinary flavanol in EPIC Norfolk (difference in log2 biomarker between bottom and top quartile of respective marker, adjusted by age, sex and BMI; 95% confidence interval and Wald-test).

|  | SREM<sub>B</sub> |  | gVLM<sub>B</sub> |  |
| --- | --- | --- | --- | --- |
|  | β (95% CI) | p | β (95% CI) | p |
| Fruits | -0.06 (-0.11; -0.02) | 0.0107 | -0.15 (-0.21; -0.1) | <0.001 |
| Vegetables | -0.09 (-0.13; -0.06) | <0.001 | -0.1 (-0.14; -0.05) | <0.001 |
| Fruits & Vegetables | -0.08 (-0.12; -0.05) | <0.001 | -0.14 (-0.19; -0.1) | <0.001 |
| Vitamin C | -0.14 (-0.17; -0.1) | <0.001 | -0.09 (-0.13; -0.04) | <0.001 |
| Tea | 0.61 (0.58; 0.65) | <0.001 | 0.28 (0.23; 0.33) | <0.001 |

**Supplemental Table 4:** List of twenty most frequency consumed fruits and vegetables in NHANES 2017-2019. Portion size was reported according to 21 CFR § 101.12.

| Food | Frequency<br>(number of mentions in dietary recall) | Portion size<br>(g) |
| --- | --- | --- |
| Banana, raw | 1112 | 140 |
| Apple [Dessert], whole, raw | 852 | 140 |
| Tomato, whole, raw | 705 | 85 |
| Grape [Black] | 463 | 140 |
| Grape [Green] | 463 | 140 |
| Orange [Blond] | 450 | 140 |
| Carrot, raw | 394 | 85 |
| Strawberry, raw | 386 | 140 |
| Cucumber, raw | 368 | 85 |
| Green bean, raw | 290 | 85 |
| Melon | 243 | 140 |
| Avocado, raw | 188 | 50 |
| Peanut | 185 | 30 |
| Almond | 172 | 30 |
| Highbush blueberry, raw | 152 | 140 |
| Pineapple | 149 | 140 |
| Lettuce [Green], raw | 126 | 85 |
| Mango | 108 | 140 |
| Grape, raisin | 101 | 40 |
| Pear, whole | 99 | 140 |

**Supplemental Figure 1:**
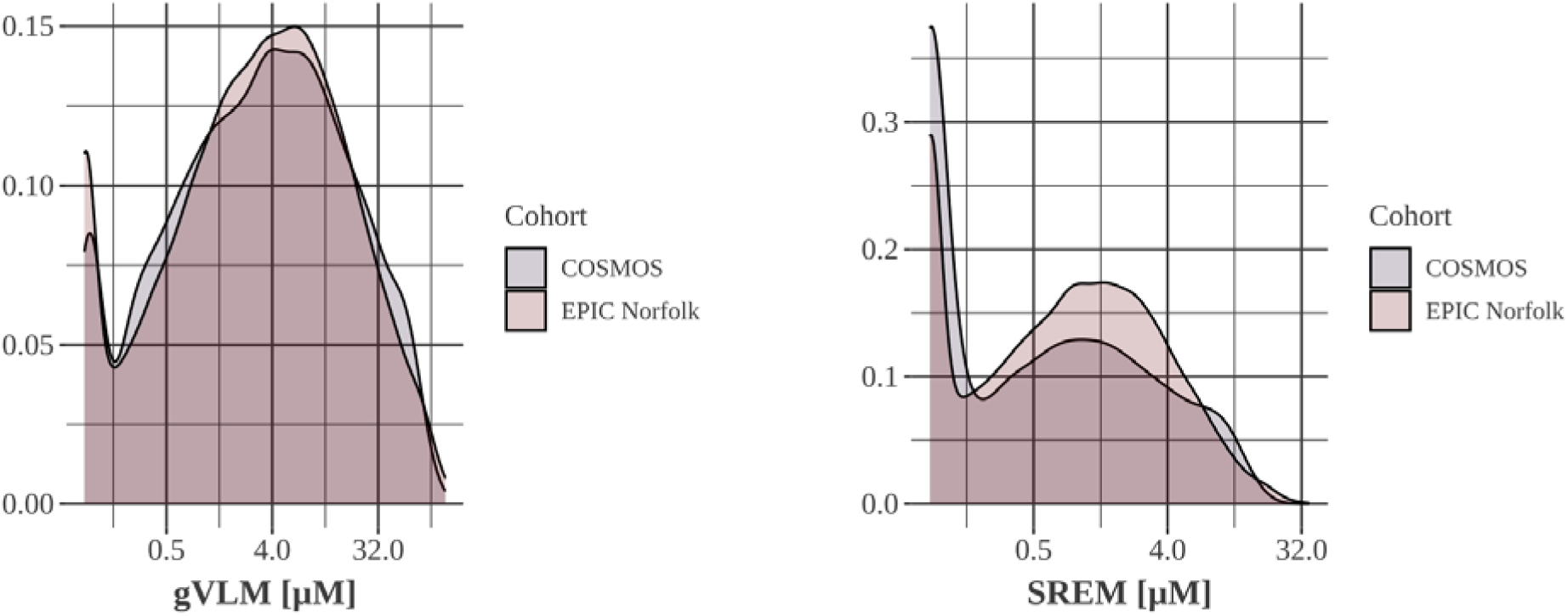
Distribution of gVLM_B_ and SREM_B_ concentration between COSMOS and EPIC Norfolk. gVLM_B_: 5-(3□,4□-dihydroxyphenyl)-γ-valerolactone metabolites; SREM_B_: structurally related (–)-epicatechin metabolites

## References

Bhagwat, S.A., Haytowitz, D.B., Prior, R.L., Gu, L., 2004. USDA database for proanthocyanidin content of selected foods [WWW Document].

Bhagwat, S.A., Haytowitz, D.B., Wasswa-Kintu, S.I., Pehrsson, P.R., 2015. Process of formulating USDA’s Expanded Flavonoid Database for the Assessment of Dietary intakes: a new tool for epidemiological research. The British journal of nutrition 114, 472–480. 10.1017/S0007114515001580

Bingham, S.A., Luben, R.N., Welch, A.A., Low, Y.-L., Khaw, K.-T., Wareham, N.J., Day, N.E., 2008. Associations between dietary methods and biomarkers, and between fruits and vegetables and risk of ischaemic heart disease, in the EPIC Norfolk Cohort Study. International Journal of Epidemiology 37, 978–987. 10.1093/ije/dyn111

Brickman, A.M., Yeung, L.K., Alschuler, D.M., Ottaviani, J.I., Kuhnle, G.G.C., Sloan, R.P., Luttmann-Gibson, H., Copeland, T., Schroeter, H., Sesso, H.D., Manson, J.E., Wall, M., Small, S.A., 2023. Dietary flavanols restore hippocampal-dependent memory in older adults with lower diet quality and lower habitual flavanol consumption. Proc Natl Acad Sci U S A 120, e2216932120. 10.1073/pnas.2216932120

Crowe-White, K.M., Evans, L.W., Kuhnle, G.G.C., Milenkovic, D., Stote, K., Wallace, T., Handu, D., Senkus, K.E., 2022. Flavan-3-ols and Cardiometabolic Health: a Guideline Recommendation by the Academy of Nutrition and Dietetics. Adv Nutr. 10.1093/advances/nmac105

Day, N., Oakes, S., Luben, R., Khaw, K.-T., Bingham, S.A., Welch, A.A., Wareham, N.J., 1999. EPIC-Norfolk: study design and characteristics of the cohort. European Prospective Investigation of Cancer. British Journal of Cancer 80 Suppl 1, 95–103. 10.2337/diacare.23.6.726

Erdman, J.W., 2023. Health and nutrition beyond essential nutrients: The evolution of the bioactives concept for human health. Mol Aspects Med 89, 101116. 10.1016/j.mam.2022.101116

Feskanich, D., Rimm, E.B., Giovannucci, E.L., Colditz, G.A., Stampfer, M.J., Litin, L.B., Willett, W.C., 1993. Reproducibility and validity of food intake measurements from a semiquantitative food frequency questionnaire. YJADA 93, 790–796.

Gaine, P.C., Balentine, D.A., Erdman, J.W., Dwyer, J.T., Ellwood, K.C., Hu, F.B., Russell, R.M., 2013. Are Dietary Bioactives Ready for Recommended Intakes?12. Adv Nutr 4, 539–541. 10.3945/an.113.004226

Harrell Jr, F.E., 2017. rms: Regression Modeling Strategies.

Khaw, K.-T., Bingham, S.A., Welch, A.A., Luben, R., Wareham, N.J., Oakes, S., Day, N., 2001. Relation between plasma ascorbic acid and mortality in men and women in EPIC-Norfolk prospective study: a prospective population study. Lancet 357, 657–663.

Kuhnle, G.G., 2018. Nutrition epidemiology of flavan-3-ols: The known unknowns. Molecular Aspects of Medicine 61, 2–11.

Lupton, J.R., Atkinson, S.A., Chang, N., Fraga, C.G., Levy, J., Messina, M., Richardson, D.P., van Ommen, B., Yang, Y., Griffiths, J.C., Hathcock, J., 2014. Exploring the benefits and challenges of establishing a DRI-like process for bioactives. Eur J Nutr 53 Suppl 1, 1–9. 10.1007/s00394-014-0666-3

McCullough, M.L., Feskanich, D., Stampfer, M.J., Giovannucci, E.L., Rimm, E.B., Hu, F.B., Spiegelman, D., Hunter, D.J., Colditz, G.A., Willett, W.C., 2002. Diet quality and major chronic disease risk in men and women: moving toward improved dietary guidance. Am J Clin Nutr 76, 1261–1271. 10.1093/ajcn/76.6.1261

Ottaviani, J.I., Borges, G., Momma, T.Y., Spencer, J.P.E., Keen, C.L., Crozier, A., Schroeter, H., 2016. The metabolome of [2-14C](−)-epicatechin in humans: implications for the assessment of efficacy, safety, and mechanisms of action of polyphenolic bioactives. Scientific reports 6, 1–10. 10.1038/srep29034

Ottaviani, J.I., Britten, A., Lucarelli, D., Luben, R., Mulligan, A.A., Lentjes, M.A., Fong, R., Gray, N., Grace, P.B., Mawson, D.H., Tym, A., Wierzbicki, A., Forouhi, N.G., Khaw, K.-T., Schroeter, H., Kuhnle, G.G.C., 2020. Biomarker-estimated flavan-3-ol intake is associated with lower blood pressure in cross-sectional analysis in EPIC Norfolk. Scientific reports 10, 17964.

Ottaviani, J.I., Fong, R., Kimball, J., Ensunsa, J.L., Britten, A., Lucarelli, D., Luben, R., Grace, P.B., Mawson, D.H., Tym, A., 2018. Evaluation at scale of microbiome-derived metabolites as biomarker of flavan-3-ol intake in epidemiological studies. Scientific reports 8, 1–11.

Ottaviani, J.I., Fong, R., Kimball, J., Ensunsa, J.L., Gray, N., Vogiatzoglou, A., Britten, A., Lucarelli, D., Luben, R., Grace, P.B., Mawson, D.H., Tym, A., Wierzbicki, A., Smith, A.D., Wareham, N.J., Forouhi, N.G., Khaw, K.-T., Schroeter, H., Kuhnle, G.G.C., 2019. Evaluation of (-)-epicatechin metabolites as recovery biomarker of dietary flavan-3-ol intake. Sci Rep 9, 13108. 10.1038/s41598-019-49702-z

Ottaviani, J.I., Sagi-Kiss, V., Schroeter, H., Kuhnle, G.G.C., 2024. Reliance on self-reports and estimated food composition data in nutrition research introduces significant bias that can only be addressed with biomarkers. Elife 13. 10.7554/eLife.92941

Ottaviani, J.I., Schroeter, H., Bier, D.M., Erdman, J.W., Sesso, H.D., Manson, J.E., Kuhnle, G.G.C., 2025. The overlooked impact of background diet and adherence in nutrition trials. Food Funct. 16, 5733–5743. 10.1039/D5FO01134E

Ottaviani, J.I., Schroeter, H., Kuhnle, G.G.C., 2023. Measuring the intake of dietary bioactives: Pitfalls and how to avoid them. Mol Aspects Med 89, 101139. 10.1016/j.mam.2022.101139

Public Health England, 2016. Eatwell Guide.

Raman, G., Avendano, E.E., Chen, S., Wang, J., Matson, J., Gayer, B., Novotny, J.A., Cassidy, A., 2019. Dietary intakes of flavan-3-ols and cardiometabolic health: systematic review and meta-analysis of randomized trials and prospective cohort studies. Am J Clin Nutr 110, 1067–1078. 10.1093/ajcn/nqz178

Rist, P.M., Sesso, H.D., Johnson, L.G., Aragaki, A.K., Wang, L., Rautiainen, S., Hazra, A., Tobias, D.K., LeBoff, M.S., Schroeter, H., Friedenberg, G., Copeland, T., Clar, A., Tinker, L.F., Hunt, R.P., Bassuk, S.S., Sarkissian, A., Smith, D.C., Pereira, E., Carrick, W.R., Wion, E.S., Schoenberg, J., Anderson, G.L., Manson, J.E., Cosmos Research Group, 2022. Design and baseline characteristics of participants in the COcoa Supplement and Multivitamin Outcomes Study (COSMOS). Contemp Clin Trials 116, 106728. 10.1016/j.cct.2022.106728

Rothwell, J.A., Pérez-Jiménez, J., Neveu, V., Medina-Remón, A., M’hiri, N., García-Lobato, P., Manach, C., Knox, C., Eisner, R., Wishart, D.S., Scalbert, A., 2013. Phenol-Explorer 3.0: a major update of the Phenol-Explorer database to incorporate data on the effects of food processing on polyphenol content. Database 2013, bat070–bat070. 10.1093/database/bat070

Sesso, H.D., Rist, P.M., Aragaki, A.K., Rautiainen, S., Johnson, L.G., Friedenberg, G., Copeland, T., Clar, A., Mora, S., Moorthy, M.V., Sarkissian, A., Wactawski-Wende, J., Tinker, L.F., Carrick, W.R., Anderson, G.L., Manson, J.E., Cosmos Research Group, 2022. Multivitamins in the prevention of cancer and cardiovascular disease: the COcoa Supplement and Multivitamin Outcomes Study (COSMOS) randomized clinical trial. Am J Clin Nutr 115, 1501–1510. 10.1093/ajcn/nqac056

U.S. Department of Agriculture, U. S. Department of Health and Human Services, 2020. Dietary Guidelines for Americans, 2020-2025.

Vogiatzoglou, A., Mulligan, A.A., Bhaniani, A., Lentjes, M., McTaggart, A., Luben, R.N., Heiss, C., Kelm, M., Merx, M.W., Spencer, J.P.E., Schroeter, H., Khaw, K.-T., Kuhnle, G.G., 2015. Associations between flavan-3-ol intake and CVD risk in the Norfolk cohort of the European Prospective Investigation into Cancer (EPIC-Norfolk). FREE RADICAL BIOLOGY AND MEDICINE 84, 1–10.

Vogiatzoglou, A., Mulligan, A.A., Luben, R.N., Lentjes, M.A., Heiss, C., Kelm, M., Merx, M.W., Spencer, J.P., Schroeter, H., Kuhnle, G.G., 2014. Assessment of the dietary intake of total flavan-3-ols, monomeric flavan-3-ols, proanthocyanidins and theaflavins in the European Union. BRITISH JOURNAL OF NUTRITION 111, 1463–1473.

Welch, A.A., McTaggart, A., Mulligan, A.A., Luben, R., Walker, N., Khaw, K.-T., Day, N.E., Bingham, S.A., 2001. DINER (Data Into Nutrients for Epidemiological Research) - a new data-entry program for nutritional analysis in the EPIC-Norfolk cohort and the 7-day diary method. Public Health Nutrition 4, 1253–1265.

White, I.R., Royston, P., 2009. Imputing missing covariate values for the Cox model. Statistics in medicine 28, 1982–1998. 10.1002/sim.3618

Wickham, H., 2009. ggplot2: Elegant Graphics for Data Analysis. Springer-Verlag New York.

Wilkinson, B.G., Perring, M.A., 1961. Variation in mineral composition of Cox’s Orange Pippin apples. Journal of the science of food and agriculture 12, 74–80. 10.1002/jsfa.2740120114

Wu, W., Zhu, Q., Wang, W., Grierson, D., Yin, X., 2022. Molecular basis of the formation and removal of fruit astringency. Food Chemistry 372, 131234. 10.1016/j.foodchem.2021.131234

Yates, A.A., Dwyer, J.T., Erdman, J.W., King, J.C., Lyle, B.J., Schneeman, B.O., Weaver, C.M., serving as an ad hoc Working Group on a Framework for Developing Recommended Intakes for Dietary Bioactives, 2021. Perspective: Framework for Developing Recommended Intakes of Bioactive Dietary Substances. Adv Nutr 12, 1087–1099. 10.1093/advances/nmab044

Yoshida, K., Bohn, J., 2015. tableone: Create “Table 1” to Describe Baseline Characteristics.

